# Expecting the Unexpected: Predicting Panic Attacks from Mood and Twitter

**DOI:** 10.1101/2023.01.26.23285057

**Authors:** Ellen W. McGinnis, Shania Lunna, Isabel Berman, Skylar Bagdon, Genevieve Lewis, Michael Arnold, Christopher M. Danforth, Peter Sheridan Dodds, Matthew Price, William E. Copeland, Ryan S. McGinnis

## Abstract

**Background:** Panic attacks are an impairing mental health problem that affects about one in 10 US adults every year. Current DSM criteria describe panic attacks as unexpected, occurring without warning or triggering events. The unexpected nature of panic attacks not only leads to increased anxiety for the individual but has also made panic attacks particularly challenging to study. However, recent evidence suggests that individuals who experience such attacks could identify attack triggers.

**Objective:** We aimed to explore both retrospectively and prospectively, qualitative, and quantitative factors associated with the onset of panic attacks.

**Method:** We remotely recruited a diverse sample of 87 individuals who regularly experienced panic attacks from 30 states in the US. Participants responded to daily questions relating to their panic attacks and wellness behaviors each day for 28 days. We also considered daily community level factors captured by the Hedonometer, a metric which estimates population-level happiness daily using a random 10% of all public tweets.

**Results:** Consistent with our prior work, most participants (95%) were able to retrospectively identify a trigger for their attack. Worse individual mood was associated with greater likelihood of experiencing a *same-day* panic attack over and above other individual wellness factors. Worse individually reported mood and state-based population level mood as indicated by the Hedonometer were associated with greater likelihood of *next-day* panic attack.

**Conclusions:** These promising results suggest that individuals who experience panic attacks may be able to expect the unexpected. The importance of individual and state-based population level mood in panic attack risk could be used to ultimately inform future prevention and intervention efforts.

## Introduction

Over 28% of US adults have experienced at least one panic attack in their lifetime, with over 11% having experienced a panic attack in the past year [1]. Panic attacks are impairing, both physically and emotionally, with physiological and psychological symptoms including increased heart rate, hyperventilation, shaking, feelings of faintness, as well as self-perceived lack of control, and fear of another panic attack occurring [2]. The DSM-5 currently defines panic attacks as an “ abrupt surge of intense fear or discomfort that reaches a peak within minutes” that comes out of nowhere [2], suggesting there are no triggers for an attack. One report on the criteria of panic attacks goes as far as to state that, “ A hallmark feature of panic disorder is that attacks occur without warning.^”^ [3]. If a hallmark feature is a lack of a trigger, then panic attacks would be almost impossible to predict, leaving those who experience panic attacks feeling a loss of control over their own bodies. However, some data suggests that many panic attacks may indeed have specific triggers [4]. If triggers of panic attacks could be identified, they may be explored further to examine whether they could predict panic attack onset, and ultimately be used to give back some semblance of control to those who experience panic attacks.

Many studies examining panic attacks in a laboratory induce panic attacks medically [5]) or experientially via interoceptive therapy [6]. We have only recently started to identify triggers of naturally occurring panic attacks [7]. The DSM-5 mentions some cognitive risk factors associated with panic attacks such as negative affect and anxiety sensitivity, but new evidence suggests anxiety sensitivity could have a causal role in panic [8]. Our previous work demonstrates nearly all (98%) of 85 participants suffering from panic attacks could identify a trigger of their latest attack [4]. Health was the most frequently identified trigger (38%), followed by conflict (19%), performance (17%), and workload (15%), with only 2% reporting they were unsure of the trigger [4]. However, participants were given a forced choice of 10 possible triggers during their panic attack, which may have impacted their responses. Trigger prevalence may have been impacted by the timing of the study as it began in March 2020, co-occurring with the start of COVID-19 pandemic related lockdowns in the United States, likely making health a more commonly identified trigger than in other years. Given the prevalence of self-reported triggers in this lone, forced-choice study, it is important to further examine panic attack triggers in an open response format outside of transient contextual factors like pandemic lockdowns to confirm these findings. Additionally, it would be beneficial to examine prospective factors that may lead to panic attacks and to investigate if it is possible to predict their occurrence.

While there is a dearth of literature on panic attack triggers outside of a laboratory, there are more studies examining triggers of acute anxiety or mood more generally. For instance, acute anxiety has been associated with prior lack of sleep [9] and caffeine use [10]. Daily mood has been associated with getting an adequate amount of sleep (8+ hours), exercise (30+ minutes), servings of fruits and vegetables (4+), water (4+ glasses) and singing/playing an instrument, as well as the cumulation of these wellness behaviors in college students [11]. Our previous study on panic attacks suggests there may be an association with wellness behaviors, such that on days that people had panic attacks, they retrospectively reported themselves as experiencing more substance use, worse eating habits, worse exercise, worse sleep quality and more stress than typical for them [4]. However, retrospective reporting is likely influenced by the occurrence and negative experience of having a panic attack, especially for highly anxious individuals [12]. In the few prospective studies on panic attacks, one found that caffeine intake was likely linked to panic attack onset as individuals with panic disorder who were given caffeine equivalent of about 4 cups of coffee in a lab setting were more likely to have a panic attack in the lab than those not given caffeine [13]. Another study followed individuals with panic disorder prospectively for 2 days using an array of physiological sensors. They found significant changes in heart rate occurred 45 minutes prior to the onset of a panic attack [14]. Therefore, there may be within-person changes prior to the onset of panic attacks that could be identified if studied longitudinally in a sample suffering from frequent panic attacks.

Beyond within-person associations of acute anxiety, mood, and panic, there is also evidence suggesting that some collective environmental factors are associated with these states. For instance, in our previous paper, health was listed as a trigger of panic attacks by 38% of respondents during the beginning of the 2020 pandemic lockdown in the United States [4]. Other studies have shown an increase of anxiety due to an increase of mortality-related thoughts due to the pandemic [15]. Collective negative events outside of the pandemic such as acts of terrorism, human losses, and political rulings affecting civil rights have been seen to impact individuals’ mental health [16,17], and specifically, anxiety levels [17], with proximity to the event, in terms of location or personal relevance, moderating the strength of that impact [17]. This connection between community-level negative events and anxiety may be even more relevant for highly anxious individuals who attend more to potential external threats [18]. The Hedonometer [19][20] is a public access tool that codes Twitter valence as a barometer for collective English-language and US state-level moods. For instance, mood based on the positive and negative valence of words used in Tweets was significantly reflective of the relative mental and physical health of those in the geographic region being studied, as well as collective negative experiences such as unemployment [21], education levels and obesity rates [19]. This metric may provide an important external measure of mood that reflects an individual’s community and relevant events that may be contributing to their daily state of anxiety and risk for panic attacks.

In the current study, we aimed to identify factors associated with the onset of panic attacks to ultimately inform future prevention and intervention efforts. We first assess qualitative, retrospective reports of panic attack triggers to examine their content and relative prevalence. We next test whether daily individual- and community-level factors are associated with a greater likelihood of experiencing a panic attack *the same-day* and/or *the next-day*.

## Method

### Procedure

Participants were recruited through advertisements on Facebook over 72 hours in Spring 2022. Ads were targeted at users identified by Facebook as “ highly anxious” and “ interest in Apple Watches.” Study eligibility required that all participants 1) lived in the United States, 2) owned an iPhone and 3) an Apple Watch, 4) were at least 18 years old, and 5) had experienced at least one panic attack in the last seven days. The protocol was approved by the University of Vermont Institutional Review Board. After e-consenting, participants were asked to complete an initial survey and then daily survey for the following 28 days. Participants earned a $20 Amazon gift card each week they were enrolled in the study.

### Measures

The *initial survey* (15 minutes) included items on basic demographic information, mental health history, panic attack history, and about possible panic attack interventions they might have used. The *daily survey* (5 minutes) included items on whether they had experienced a panic attack and associated descriptive information including what they believed their trigger was (open text response), their mood, sleep quality, eating habits (each on a scale of 1-poor, 2-fine, 3-neutral, 4-good, 5-great), stress (on a scale of 1-low stress to 5-very high stress), conflict (yes or no), exercise minutes (open numeric response), and substance use inquiring separately about caffeine, alcohol, prescription drugs, recreational drugs (yes or no) (see Supplemental Table 1 for more detail). Participants were also asked to upload their Apple Watch data weekly after wearing it every day and to use an app, PanicMechanic^™^ to record their heart rates during any panic attack they experienced [4,22,23]. Only data from surveys are included in subsequent analyses.

The Hedonometer is a sentiment analysis technique that measures happiness demonstrated in an extensive collection of texts [24]. It assigns sentiment scores to individual words using Amazon’s Mechanical Turk online marketplace and is based on a wordlist of 10,222 unique words compiled by merging the most frequent 50,000 words rated by from four disparate corpora, Twitter, Google Books, the New York Times and music lyrics [20]. The happiness score ranges from 1 (least happy) to 9 (most happy). The Hedonometer extracts word frequency, assigns happiness scores to each word, and computes the weighted average happiness of a given text [20]. This tool can be used on Twitter data to characterize the overall happiness of a group of geographically collocated people (e.g. [25]). Here, we characterize English language Tweets and US state Tweets as levels of daily mood for the duration of the study to inform our analyses.

### Sample

After completing an eligibility questionnaire, 107 individuals signed an e-consent form. Twenty of those participants did not complete any study questionnaires besides the e-consent and could not be reached for follow-up. Eighty-seven participants completed at least one day of “ daily questionnaires” and are included in the subsequent analyses. There was significant attrition in this completely remote study. At the end of each week, participants were withdrawn by the study team if they had completed less than 15% of their weekly surveys. Of the 87 who completed at least one day of surveys, 35 participants were withdrawn after week one, another 25 participants were withdrawn after week two, and one participant was withdrawn after week three. Thirty-nine participants completed 4 weeks of data with more than 15% adherence each week (completers). Attrition analyses revealed that completers did not significantly differ from those who were withdrawn by educational attainment, annual income, age, race/ethnicity, and mental health diagnostic status, or number of lifetime panic attacks. However, participants identifying as women were more likely to complete the four weeks of the study (55%) than those identifying as men (29%)(*x* (1) = 5.21, *p* = .022). All 87 participants were included in analyses, completing a combined total of 1247 days of daily questionnaires.

### Analytical Plan

Descriptive analyses were conducted on the initial survey responses to characterize our study sample including demographics, mental health, and panic attack information.

Qualitative content analysis [4,26] was conducted on open text responses of panic attack triggers. Two coders read all responses and developed categories with respective definitions based on commonalities amongst responses. Categories were then discussed by coders together and 16 categories were determined via consensus. The two coders then independently coded each response to fit into one of the 16 categories. Multiple responses were coded from a single respondent if it included the word “ and” or a comma. Any discrepancies between coders were sent to a third coder to reach consensus.

Mixed Regressions, with an autoregressive covariance structure were used to estimate the prevalence of a panic attack. In this approach, participant ID is introduced as a cluster (class) variable to account for repeated, correlated observations within individuals. Robust variance estimates (i.e., sandwich type estimates) adjusted the standard errors of the parameter estimates for the within-person nesting of observations. The alpha value for significance testing was set at <.05. The number of days since the start of the study was included as a covariate in all analytical models. All statistical analyses were performed in SPSS (Version 28.0.1.1, IBM). Regression analysis began by examining demographic variables in univariate models. Any demographic variables found to be significantly associated with panic attack likelihood during the study were included as covariates in subsequent models. Individual-level mood scores and wellness behaviors and community-level (Hedonometer) mood scores were then examined using univariate models for outcomes of same-day panic attack occurrence and next-day panic attack occurrence using lag sequential analyses. Finally, any individual-level or community-level factors found to be significantly associated with likelihood of having a panic attack in univariate models were included in simultaneous models for same-day and next-day panic attack occurrence.

## Results

### Sample description

Of the 87 participants with at least one day of data (see **Table 1**), the majority (63.2%) identified as women, and the average age was 36.07 (SD: 10.83). The sample is racially/ethnically diverse with about 50% identifying as a racial minority, and geographically diverse, as participants reported living across 30 states. Almost two-thirds of participants have a bachelor’s degree or graduate degree and the majority of participants earned over $75,000 annually. Over half of participants have at least one mental health diagnosis, the most common being generalized anxiety disorder (GAD) and major depressive disorder (MDD). Most participants in the study had tried at least one intervention for their panic attacks, with the most common being medication and self-care (i.e, meditation, exercise). The mean number of panic attacks in the month prior to recruitment was 4.34 (SD=5.36), and participants reported an average of 72.9 (SD=187.28) panic attacks across their lifetime. Over the 28 days of the study, there were 268 panic attacks reported by 64 unique people. The average reported length of a panic attack was 17.58 minutes (*SD=* 16.7, R=1-120 minutes), and the mean reported severity of the primary panic attack was 6.62 (*SD*=1.7) out of a 1-10 rating (10 being “ the most anxious I have ever felt”).

**Table 1.** Demographic Description of Sample.

| Characteristic | Details | % (*n*) |
| --- | --- | --- |
| Gender | Woman | 63.2% (55) |
|  | Man | 35.6% (31) |
| Race | White, Non-Hispanic | 49.4% (43) |
|  | African American | 33.3% (29) |
|  | Asian-Pacific | 8.0% (7) |
|  | Latinx | 4.6% (4) |
|  | Bi-racial | 3.4% (3) |
| Education | High school or GED | 8.0% (7) |
|  | Some college | 25.3% (22) |
|  | Bachelor's degree | 40.2% (35) |
|  | Graduate degree | 25.3% (22) |
| Income | \$0-\$25,000 | 16% (14) |
| | \$25,001-\$50,000 | 13.6% (12) |
| | \$50,001-\$75,000 | 11.3% (10) |
| | \$75,001-\$100,000 | 29.8% (26) |
| | >\$100,000 | 27.6% (24) |
| Mental Health | At least 1 diagnosis | 53.1% (51) |
|  | GAD | 60.8% (31) |
|  | MDD | 45.1% (23) |
|  | PTSD | 17.7% (9) |
|  | Panic Disorder | 9.8% (5) |
|  | Other | 33.3% (17) |
| Interventions | None | 17.2% (15) |
|  | Prescription Medication | 54.0% (47) |
|  | Therapy | 29.9% (26) |
|  | Emergency Department | 12.6% (11) |
|  | Self-Help (meditation) | 60.9% (53) |

### Retrospective qualitative panic attack triggers

Of the 268 panic attacks, the triggers most reported (in total and among different participants) were related to emotional health (see **Table 2**). Other commonly reported triggers (in order of reporting) were related to relational conflict, physical health, physical environment, social situations, sleep, work and school, finances, substances, and thoughts. Few people (≤3) reported panic from concern for others, loss, being confronted with bad news, and therapy. Three individuals reported being unsure about the cause of their panic attack. All participants provided a response to this question.

**Table 2.** Frequencies of Self-Reported Triggers.

| Category | Details | PA Freq<br>(N=268) | Part Freq<br>(N=64) | Example Triggers |
| --- | --- | --- | --- | --- |
| Emotional Health | Anxiety, depression, PTSD, stress, overwhelmed, worry, anger, fear | 96<br>(35.82%) | 31<br>(29.69%) | Feeling overwhelmed |
| Relational Conflict | Arguments, break ups/divorce, family issues, conflict with others | 41<br>(15.30%) | 24<br>(37.5%) | I had a fight with my husband |
| Physical Health | Cancer diagnosis, surgery, catheter, hospitalization, falling, accidents | 38<br>(14.18%) | 19<br>(29.69%) | Headache |
| Physical Environment | Sounds, sensations, environment, driving, airplanes, travel | 28<br>(10.45%) | 18<br>(28.13%) | Being in a car while it's snowing |
| Social Situation | Social anxiety, public, social issues | 19<br>(7.09%) | 13<br>(20.31%) | too many people |
| Sleep | Nightmares, lack of sleep, sleep issues | 16<br>(5.97%) | 12<br>(18.75%) | Lack of sleep |
| Finances | Bills, finances, broken car, lost job, financial stress | 15<br>(5.60%) | 7<br>(10.94%) | Financial stress |
| Substances | Triggers related to substances (alcohol/caffeine/drug use) | 15<br>(5.60%) | 6<br>(9.38%) | Some drinks |
| Work/School | Issues with work or school | 14<br>(5.22%) | 8<br>(12.5%) | Nervous about exam tomorrow |
| Thoughts | Overthinking, busy thoughts, intrusive | 11<br>(4.10%) | 5<br>(7.81%) | Deep thoughts |
| Concern | Concern for other's health | 6<br>(2.24%) | 6<br>(9.38%) | My little boy fell off the stairs |
| Bad news | Friendship loss or death | 3<br>(1.12%) | 3<br>(4.69%) | Bad news |
| Loss | Words did not portray meaning enough to code into category | 3<br>(1.12%) | 3<br>(4.69%) | Death of loved one |
| Therapy | During or because of therapy | 3<br>(1.12%) | 1<br>(1.56%) | During therapy |
| Unsure | Not knowing the trigger | 7<br>(2.61%) | 3<br>(4.69%) | Unsure |
| Too vague | Words did not portray meaning enough to code into category | 3<br>(1.12%) | 3<br>(4.69%) | Personal situation |

### Associations between demographics and panic attacks

In univariate models of demographic factors, male gender *(b*=.14, se=.045, *p*=.003), minority race *(b*=-.10, se=.044, *p*=.026), and no lifetime diagnosis of a mental health disorder *(b*=.12, se=.042, *p*=.006) were each significantly associated with greater likelihood of experiencing a panic attack, thus were included as covariates in subsequent models. Age *(b*=-.003, se=.002, *p*=.179), income *(b*=-.005, se=.003, *p*=.098), education *(b*=-.03, se=.025, *p*=.267), and whether they tried interventions to reduce their panic attacks in the past *(b*=.08, se=.055, *p*=.156) were not significant.

### Retrospective reports of associations between daily behaviors and same-day panic attacks

For same-day panic attacks 1247 days were analyzed (268 panic days wherein a panic attack was experienced) (see **Table 3**). Univariate models demonstrated that after adjusting for gender, race, mental health status, and day in study, several wellness behaviors were associated with greater likelihood of experiencing a panic attack that day including mood, sleep quality, eating quality, stress, caffeine intake, recreational drugs, having a conflict, and it being a weekend. Hedonometer scores were not related to same-day panic attack likelihood. In a simultaneous model, only mood and weekend remained significant such that worse mood and it being a weekend day (Saturday or Sunday) were related to greater likelihood of a panic attack.

**Table 3.** Same-day panic attack likelihood association with wellness behaviors and hedonometer values in univariate and simultaneous models.

|  | Univariate Models |  |  | Simultaneous Model |  |  |
| --- | --- | --- | --- | --- | --- | --- |
| Daily Individual Items | OR | 95% CI | p | OR | 95% CI | p |
| Mood | 0.88 | 0.86-0.90 | <.001 | 0.97 | 0.94-0.99 | <.001 |
| Sleep Quality | 0.91 | 0.89-0.93 | <.001 | 1.01 | 0.98-1.03 | .099 |
| Eat Quality | 0.90 | 0.89-0.92 | <.001 | 1.00 | 0.97-1.03 | .731 |
| Stress | 1.14 | 1.12-1.16 | <.001 | 1.00 | 0.97-1.02 | .691 |
| Exercise Y/N | 1.05 | 0.99-1.11 | .121 | -- | -- | -- |
| Caffeine Y/N | 0.92 | 0.87-0.98 | .005 | 1.01 | 0.95-1.07 | .104 |
| Alcohol Y/N | 0.94 | 0.87-1.03 | .186 | -- | -- | -- |
| Recreational Drugs y/n | 0.80 | 0.68-0.94 | .006 | 1.10 | 0.93-1.31 | . |
| Prescription Drugs y/n | 1.04 | 0.89-1.03 | .233 | -- | -- | -- |
| Conflict y/n | 0.82 | 0.76-0.88 | <.001 | 1.03 | 0.95-1.11 | .442 |
| Weekend y/n | 0.47 | 0.43-0.51 | <.001 | 0.93 | 0.83-1.04 | .197 |
| Daily Community Items |  |  |  |  |  |  |
| English Language Mood | 0.76 | 0.23-2.48 | .651 | -- | -- | -- |
| State Mood | 0.44 | 0.16-1.19 | .106 | -- | -- | -- |

Note: Day in study and Gender, Race and Mental Health as covariates in each model

### Prospective reports of associations between daily behaviors and next-day panic attacks

For next-day panic attacks 1160 days were analyzed (250 panic days wherein a panic attack was experienced) (see **Table 4**). Univariate models demonstrate that adjusting for gender, race, mental health status, and day in study, only worse individual-level mood, and worse Hedonometer measured state-level mood were significantly associated with greater likelihood of a next-day panic attack. Both individual and state-level mood remained significant in a simultaneous model, demonstrating unique variance from each factor.

**Table 4.**
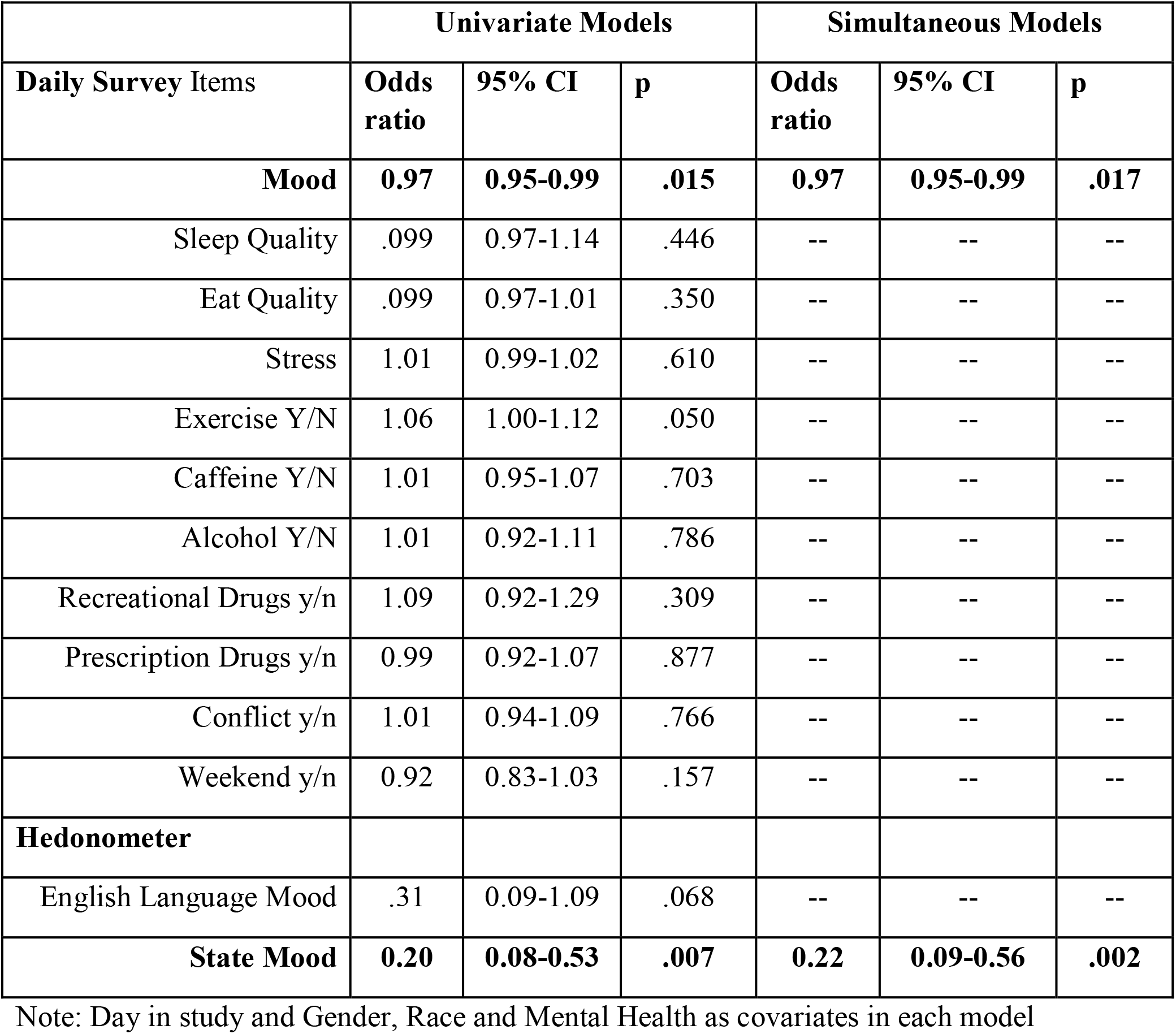
Next-day panic attack likelihood association with wellness behaviors and hedonometer values in univariate and simultaneous models.

For further examination of significant trends, State Mood was binned according to Individual Mood grouping percentiles. For example, 10% of days were self-reported as a 5 (Great) on mood, thus the top 10% of Hedonometer scores were binned as a 5. Individual and binned State Mood are visible in **Figure 1**. For Individual Mood, post-hoc analyses inclusive of previous model covariates revealed that having a 5 (“ Great”) mood rating was associated with lesser likelihood (about 8.5 times less likely) of having a next-day panic attack than each other mood rating (1(“ Poor”), 2 (“ Fine”), 3 (“ Neutral”), and 4 (“ Good”)). Additionally, having a 4 (“ Good”) mood rating was associated with lesser likelihood (about 1.5 times less likely) of having a next-day panic attack than a 1 (“ Poor”) or 2 (“ Fine”) mood rating. For State Mood, a state scoring of 1 (binned to reflect an Individual Mood rating of “ Poor”) was associated with a *greater* likelihood (about twice as likely) of having a next-day panic attack than any other score (3, 4, and 5). Additionally, having a 2 (binned to reflect an Individual Mood rating of “ Fine”) mood score was associated with a *greater* likelihood (about 1.5 times as likely) of having a next-day panic attack than a 3, 4, or 5 mood score. Thus, it seems that experiencing a “ Great” individual mood was protective against having a panic attack and living in a state with a daily “ Poor” population-level mood was a risk for having a panic attack the next day.

**Figure 1.**
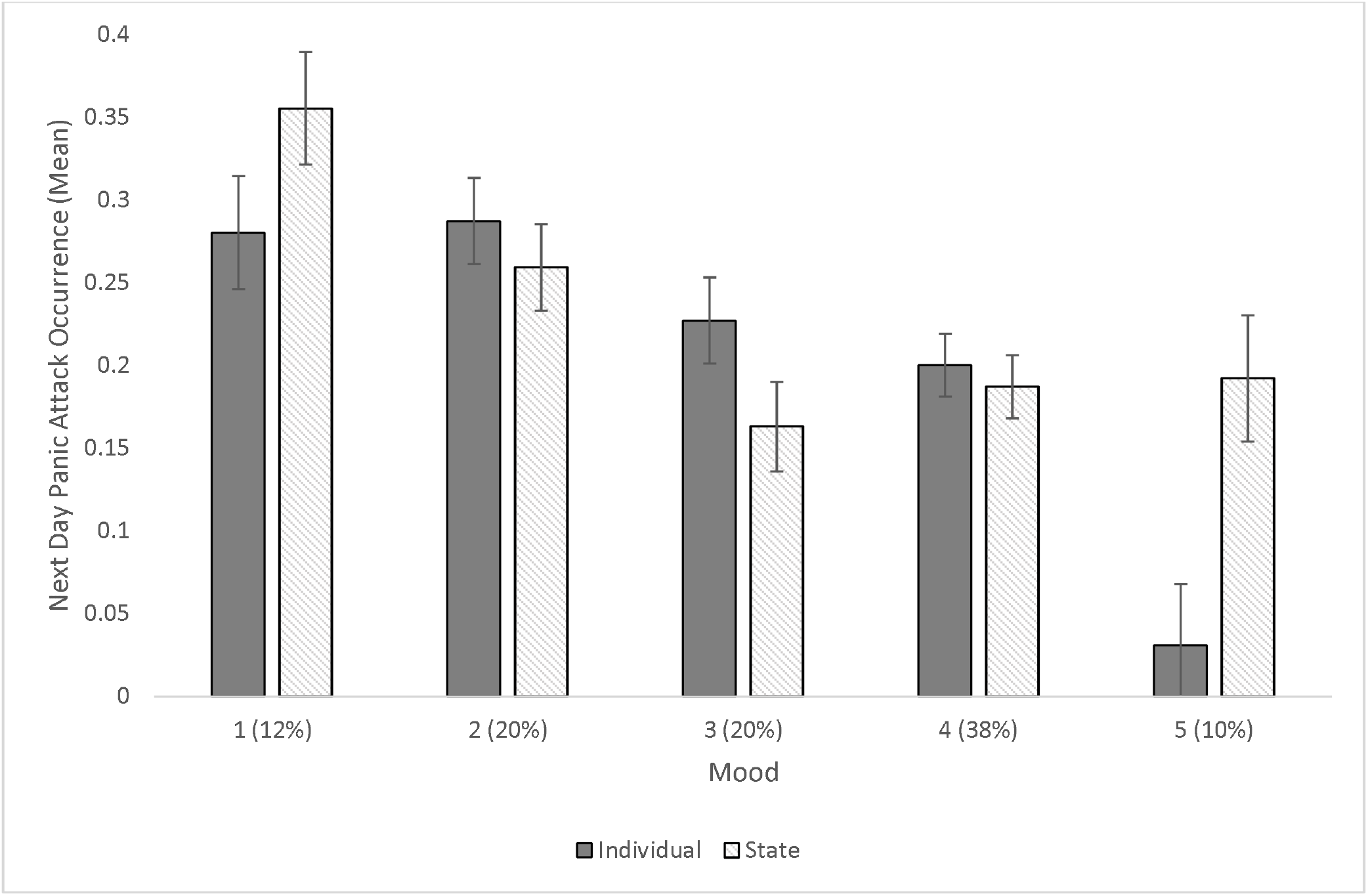
Individual Level Mood and Community Level State Mood on Next Day Panic Attack Occurrence Mean Controlling for Day in Study, Participant Race, Gender, and Mental Health Status. Note: Individual Mood is based on self-reported ratings of 1 (Poor) to 5 (Great). State Mood is indicative of State Hedonometer score, binned according to Individual Mood grouping percentiles. For example, 10% of days were self-reported as a 5 (Great) on mood, thus the top 10% of Hedonometer scores were binned as a 5.

## Discussion

The purpose of the current study was to investigate triggers of panic attacks, using both qualitative retrospective and quantitative prospective data, to explore whether panic attacks can be predicted. We found that in our sample of people who commonly experience panic attacks, most (95%) can retrospectively identify a trigger for their attack when asked. We identified factors reported retrospectively (about the past 24 hours), associated with likelihood of *same-day* panic attack such that worse individual mood, over and above other individual daily wellness factors, was associated with greater likelihood of experiencing a panic attack. Finally, we identified factors reported prospectively associated with likelihood of *next-day* panic attack such that an individual with a “ poor” mood was 8.5 times more likely to have a panic attack the next day compared to an individual with a “ great” mood, and that an individual living in a state with a “ poor” daily population-based mood (assessed via Twitter [20]) was twice as likely to have a panic attack the next day compared to an individual living in a state with a “ great” daily population-based mood.

### Retrospective qualitative trigger analysis

In contrast with DSM panic attack criteria and associated reporting describing unexpected nature and lack of trigger as hallmarks of a panic attack, we found that for most people and for most panic attacks, a trigger can be identified retrospectively. The most common trigger identified, which included about a third of panic attacks (and people), was related to emotional health. However, emotional health, including feelings of depression, anxiety and being overwhelmed, do not tend to describe specific instances of time or place. Therefore, to people identifying this common category as a trigger to their attack, the attack itself may very well be unexpected. This trigger category is consistent with the DSM, which states that negative affect and anxiety are “ risk factors” for panic attacks [3], which may indeed be a more indicative term than “ trigger” due to their non-specific nature. We need to learn more about the patterns and metrics of emotional health triggers to use them to help predict panic attack onset, such as timing between feeling onset or peak rating and panic attack onset. On the contrary, however, other reported triggers related to relational conflict, physical surroundings, social situations, substances, some work-related issues (upcoming exam), and sleep each potentially have specific onsets or timing which suggests they could be used to predict panic attacks. In terms of prevalence, relational conflict and physical health were the second and third most commonly self-reported triggers, respectively. These findings are consistent with our past research which issued a force choice option of triggers, wherein “ health” was the most common choice, and conflict was the next most common [4]. For the current analyses, we had hypothesized that health may be relatively less common than in our past study that coincidently occurred at the onset of pandemic related shutdowns. However, it appears even as pandemic-related illness and cues have decreased somewhat in our society two years later, health continues to be a top concern. Similar to our previous study, fewer than 5% of people stated “ unsure” as a panic attack trigger, which suggests that lack of prediction may *not* in fact be a hallmark of panic attacks and should be further assessed for improved recommendations on definitions and criteria.

### Retrospectively reported associations with same-day panic attack

Many correlates of same-day panic attacks were consistent with the literature on panic attacks, acute anxiety, and daily mood. For instance, retrospectively reported mood, sleep quality, eating quality, stress, caffeine intake, recreational drug intake, experiencing a relational conflict, and it being a weekend were each separately associated with having a panic attack that day. Interestingly, only mood rating remained significant in the simultaneous model over and above all these factors. Wellness behaviors have been found to be highly associated with daily mood (Copeland et al., 2022) and thus in our results, it may be that engagement in wellness behaviors are highly correlated with daily mood; which is in turn, related to panic attack likelihood. It is interesting that mood and not ‘stress’ remained significantly associated with panic attack likelihood. Mood has been defined as “ a pervasive and sustained feeling tone that is endured internally and which impacts nearly all aspects of a person’s behavior in the external world” (Sekhon & Gupta, 2022), whereas stress is an emotional state, and a response to external factors (Tsigos et al., 2020). This finding may suggest that the negative impacts of stress are only one component of mood and could possibly be mitigated by a positive mood, consistent with literature demonstrating that individuals with more positive moods have the ability and tools to adapt better to stressors (Leger, Charles & Almedia, 2019). Mood is powerful and can change the way an individual responds to stressors, such as having a panic attack.

### Prospectively reported associations of mood and next-day panic attack

Our results suggest that we can identify prospective associations between panic attacks and individual mood rating, and Hedonometer state-level mood collected the previous day. Specifically, experiencing a “ Great” mood seems to protect against the likelihood of having a panic attack. As stated above, this is consistent with previous literature demonstrating that positive moods allow individuals to better adapt to stressors (Leger, Charles & Almedia, 2019) that perhaps would have triggered a panic attack otherwise. Knowing now that mood has the potential to be a protective factor for panic attacks, efforts in developing interventions could focus on cultivating positive mindsets consistent with interventions within positive psychology [27]. For people who suffer regularly from panic, state-level mood measured via Hedonometer also had a significant impact on next-day likelihood for experiencing a panic attack. Specifically, it seems that the lowest 32% of state measured Hedonometer days increased the likelihood for experiencing a next-day panic attack. For instance, one day in the lowest scoring state mood grouping was April 12, 2022 for the state of New York, on which there was a mass shooting in a New York City subway resulting in multiple injuries. The reporting and public response to this tragedy contributed, in part, to New York’s state Hedonometer score. Events such as these were associated with the likelihood of a next-day panic attack for individuals in our study. Results demonstrated that English language Hedonometer scores exhibited only trend-level significance, suggesting that proximity (physical, and likely emotional) to the geographical location of the event effects the strength of the association between community-level mood and likelihood of experiencing a panic attack. Acknowledging that individual wellbeing is affected by societal events is consistent with previous literature [16,17][28], and in this particular case of panic attacks, demonstrates that individual physiology is acutely impacted. This association between individual physiology and societal events may be strengthened in individuals who experience regular panic attacks and as anxious individuals may pay more attention to negative occurrences in the news [29][30]. Results suggest that people who suffer from panic attacks may be able to self-intervene by intentionally distancing themselves from news outlets and social media or, spending more time on anxiety prevention practices like self-care on days with particularly high levels of societal turmoil. Perhaps just the knowledge that state level mood and events are associated with having a panic attack the next day could be helpful psychoeducation for individuals who suffer from panic attacks, as understanding of why and how panic attacks occur has been shown to be an effective intervention at least short-term [31,32]. Being alerted to a relatively low Hedonometer score day in your state could potentially allow for personalized risk management based on individual preference. These prospectively identified factors provide exciting grounds for future research investigating an algorithm to help people predict their own panic attacks.

### Limitations

This study has several potential limitations. First, there was high attrition, especially within the first two weeks. Sixty-eight percent of participants who submitted at least some daily data were withdrawn by the end of the second week due to low adherence to questionnaires.

Completing daily questionnaires can be burdensome and it is possible motivation could have been increased with more research team engagement or better incentive system based upon individual survey completion and bonuses. Furthermore, most participants in our study had achieved some level of higher education and had an income over $75,000 dollars. Therefore, we must caution the generalization of any conclusions as our sample is not representative of the United States population. Attrition and sample demographics both pose risks to the external validity of this study. Additionally, participants reported their mood, sleep, eating, and exercise habits after reporting whether they had a panic attack. It is possible and perhaps likely that the same-day factors found to be significantly associated with panic attacks were biased by the occurrence of the attack. Further research with a larger and more diverse sample size, and more accurate ways to measure physical and mental health are needed to support the data on predictors of panic attacks to inform more generalizable conclusions.

### Conclusion

Overall, we have presented promising data suggesting that individuals who experience panic attacks may be able to expect the previously unexpected. Additionally, being able to report panic attack triggers retrospectively may be the rule, not the exception. However, reported triggers only sometimes relay specific information that would be necessary to help predict panic attack onset. Prospectively measured individual and state mood could help an individual predict their risk for experiencing a panic attack the next day. This information could be very powerful as psychoeducation about *how* a panic attack occurs has been a front line of intervention [32], thus with replication and further exploration of triggers and predictive factors, adding *why* and *when* a panic attack occurs to psychoeducation intervention could support individuals who suffer from panic attacks even more effectively in the future.

## Data Availability

Data produced in the present study are available upon reasonable request to the authors for valid research purposes.

